# Theory-based determinants of modern contraceptive use in sub-Saharan Africa: an analysis of demographic and health surveys

**DOI:** 10.1101/2022.07.20.22277859

**Authors:** Raphael Adu-Gyamfi, Juliana Enos, Dorcas Obiri Yeboah, Kwasi Torpey

## Abstract

**Introduction:** Despite improved access to modern contraceptives in sub-Saharan Africa (SSA), the region has the highest fertility rate. Although modern contraceptive usage and its determinants in SSA have been assessed, most authors were not guided by behavioral change theories. This study sought to assess the modern contraceptive coverage in SSA and identify the theory-based determinants that need to be considered in demand creation interventions.

**Methods:** Data was obtained from the most recent demographic and health surveys conducted across 37 countries in SSA. Estimates of country-specific and pooled Regional modern contraceptive coverage were generated from 501,324 responses. Logistic regression was used to assess the relationship between modern contraceptive use and determinants selected based on the Health Belief and Social-Ecological behavior change models.

**Results:** Modern contraceptive coverage in SSA was 22.26% (95% CI: 17.91, 26.60). The health belief model determinants of modern contraceptive use included last birth by caesarian section (AOR=1.44, 95% CI:1.31,1.59), hearing of family planning at the health facility (AOR=1.18, 95% CI:1.12,1.24), or from at least one media source, being able to negotiate condom use (AOR=1.65, 95% CI: 1.55,1.76), and having a previous terminated pregnancy (AOR=0.76, 95%CI: 0.71, 0.81). The social ecological model determinants of modern contraceptive use included being above 24 years, having at least primary education, non-urgent need for a last child, and being involved in decision-making concerning personal health (AOR=1.81,95% CI:1.71,1.92).

**Discussion:** Modern contraceptive coverage in SSA is low. Age, educational status, past obstetric history, exposure to family planning information, ability to negotiate condom use or make personal health-related decisions, and the need for a child were the determinants for modern contraceptive use in the region. Countries need to develop context-specific interventions considering these determinants to help improve coverage and reduce the poor maternal and child health outcomes and the developmental gaps resulting from unplanned pregnancies.

## Introduction

In recent times, the rapid population growth across the globe is leading to immoderate resource use, waste production, and environmental degradation[1], and hindering the achievement of all the 17 sustainable development goals(SDG), especially in developing countries[2]. Although family planning is to help limit this growth, unintended pregnancies are still prevalent and leading to very poor outcomes across the globe[3–5]. Between 2015 and 2019, approximately 121 million unintended pregnancies occurred globally each year, with about a third of them ending in abortions[6]. Despite a general decline in unintended pregnancies since 1990, it remains high in sub-Saharan Africa (SSA) [7].

Sub-Saharan Africa (SSA) is the second most populous region with the highest fertility rate (4.4 children) and yearly increase in population (2.49%)[8,9]. Between 2013 and 2016, an average of 14 million unintended pregnancies were recorded annually[7] in the region, some of which were attributed to low socio-economic status, coerced contraceptive decision-making and failure of family planning methods[6,10–13]. To reduce the failure rates, the correct and consistent use of modern contraceptives has been recommended over the traditional ones due to their contraceptive efficacy[4,14–16].

### Determinants of modern contraceptive use

To initiate and sustain behaviors needed to prevent unwanted pregnancies, there have been several calls to use theory-driven strategies[10,14,17,18]. Although access to modern contraceptives has improved in several SSA countries since 1990, researchers assessing factors contributing to their differential use were not guided by behavioral change theories in selecting the determinants[17,18]. However, several theories have served as frameworks to predict contraceptive behavior among different populations. One of the first to be used is the Health belief model (HBM), which relies on cognitive factors oriented toward goal attainment (i.e., motivation to prevent pregnancy) [17].

The constructs of the HBM concerning modern contraceptive use include perceived vulnerability to pregnancy and its complications, perceived threats or challenges if pregnancy occurs, perceived benefits in preventing pregnancy, perceived barriers to contraception, cues to action for contraceptive use, and self-efficacy in the use of modern contraceptives[17,19].

However, the HBM is limited in its predictive abilities because it fails to account for people’s individual, economic, and environmental factors in determining decisions to adopt behaviors. To account for these factors, the Social Ecological Model (SEM) has been recommended for use together with the HBM constructs by authors in Social Psychology[17,18].

The SEM has four constructs that assess behavioral influences at the individual, interpersonal, community, and societal levels[20]. Crosby has demonstrated that age, marital and socio-economic status, level of education, and personal fertility desires greatly influence contraceptive use at the individual level[21]. At the interpersonal level, Fisseha found the power relations between the couple, the husband’s attitude, and opposition to contraceptive use to play a role, while the type of residence was influential at the community level[22]. At the societal level, women’s empowerment and access to contraceptives were noticed by Paul to determine contraceptive use [23].

Although a combination of the HBM and SEM constructs provides a comprehensive list of determinants for contraceptive behavior, their use in identifying determinants of modern contraceptive use in SSA has not been documented. Therefore, the authors sought to assess the modern contraceptive coverage and identify the theory-based determinants of their use in SSA.

## Methods

### Data Sources

Data for the study was obtained from measure DHS programme[24] after submitting a concept note. The most recent Demographic and Health Survey (DHS) from 37 countries in SSA whose data is available on the DHS website were included.

### Study Design

“DHS is a nationally representative household survey conducted at regular intervals to provide countries with updated information on different health topics such as maternal and child health, reproductive health, fertility, nutrition, mortality, and HIV/AIDS. The survey follows a two-stage stratified cluster sampling design. Large geographic areas are selected as enumeration areas(EA) in the first stage through the Probability Proportional to Size (PPS) approach, where a relatively larger EA is more likely to be selected than a smaller one. A pre-determined number of households are selected from each EA in the second stage. Eligible participants in those households (women aged 15 to 49 years and men 15 to 59 years) are then interviewed on various topics using comparable questionnaires across countries”[25]. For all 37 countries, data from Individual Recode (IR) file was used in the analysis.

### Primary outcome

The study’s outcome is modern contraceptive use, which was derived from a variable in the DHS questionnaire that assessed the type of contraceptive being used by respondents at the time of the survey. Its responses included no method, folkloric, traditional, and modern methods. Male and female sterilization, the contraceptive pill, intrauterine contraceptive device, injectable, implants, male and female condoms, the diaphragm, contraceptive foam and jelly, lactational amenorrhea, standard days method, and other country-specific modern methods aside abortion and menstrual regulation were classified as modern. The traditional ones include periodic abstinence and withdrawal methods, while the folkloric ones involve using herbs, amulets, and other spiritual approaches.

Respondents who reported using modern contraceptives were coded as ‘1’, and those using the folkloric, traditional, and no methods were coded as ‘0’.

### Independent variables

The independent variables for modern contraceptive use were selected based on four constructs from the HBM and three from SEM. Table 1 lists the constructs and their respective variables. Some of the variables were recoded to aid analysis. A new variable was created to determine the number of media sources from which respondents receive family planning information. Row totals of variables seeking to determine if respondents had heard of family planning on radio, TV, or in a newspaper were obtained for each respondent and coded as found in table 1. The “very poor” and “poor” categories in the wealth index variable, as well as the “rich” and “very rich” categories of the variable, were recoded as “poor” and “rich,” respectively. Regarding involvement in decision making concerning contraceptive use and decision making concerning the respondent’s health, the responses were coded as “involved” if the respondent took those decisions alone or with their partner and “not involved” if the decisions were made solely by the partner or other people. Details of the codes for the respective independent variables are found in table 1.

**Table 1.** Theory-based independent variables for modern contraceptive use.

| Theory | Constructs | DHS Variables | Coding |
| --- | --- | --- | --- |
| Health Belief Model | Perceived vulnerability | History of a terminated pregnancy | No<br>Yes |
|  | Perceived pregnancy threat | Last delivery by caesarian section | No<br>Yes |
|  | Cues to action | The number of media sources for FP information:<br>a) Heard of family planning on the radio<br>b) Heard of family planning on TV<br>c) Heard of family planning in the newspaper | 0<br>1<br>2<br>3 |
|  |  | Was told of family planning at the health facility | No<br>Yes |
|  | Self-efficacy | Can refuse sex | No<br>Yes<br>Do not know |
|  |  | Can ask the partner to use a condom | No<br>Yes<br>Do not know |
| Social Ecological Model | Individual | 1. Age | 15-24<br>25-34<br>35-49 |
|  |  | 2. Marital status | Married<br>Not married |
|  |  | 3. Educational level | No education<br>Primary<br>Secondary<br>higher |
|  |  | 4. Wealth index | Poor<br>Middle<br>Rich |
|  |  | 5. Need for last-child | Urgent<br>Later<br>No more |
|  | Interpersonal | Being involved in decision making concerning contraceptive use | Involved<br>Not involved |
|  |  | Being involved in decision making concerning own health | Involved<br>Not involved |
|  | Community | Type of residence | Urban<br>Rural |

### Statistical Analysis

The downloaded and merged data was cleaned using STATA/ IC version 16.1 software. Sampling weights were applied using the primary sampling units, strata and women’s individual weights to account for the complex survey design. Using the *svy linearized* command, the modern contraceptive coverage for each country was assessed by cross-tabulation between the outcome variable and the country variable to obtain row percentages. A random-effect meta-analysis using the metan stata module was conducted to generate the pooled SSA estimate for modern contraceptive coverage. The variables used included the country-specific modern contraceptive coverage, standard errors, and sample population. Subgroup analysis was also conducted using the sub-regional classification of the countries to obtain estimates for West, Central, Eastern, and Southern Africa and was presented with a forest plot(Fig 1).

**Fig 1.**
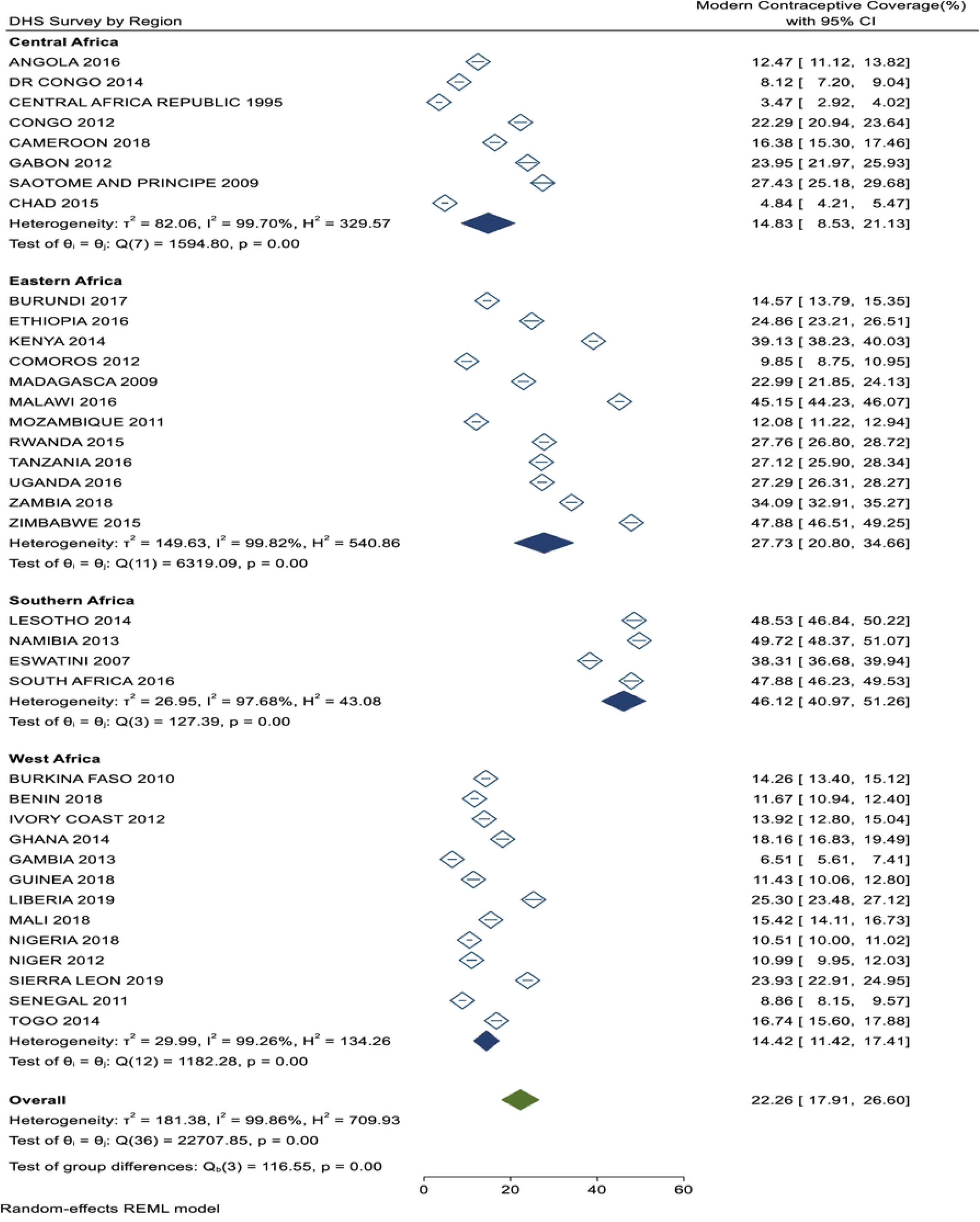
Forest plot of country-specific modern contraceptive use and pooled estimate by sub-region.

To identify the determinants of modern contraceptive use, the HBM and SEM variables, as contained in table 1, were assessed for association with the outcome variable by conducting a bi-variable multilevel regression analysis, and the statistically significant ones (p≤ 0.05) were considered for inclusion in their theory-specific logistic regression model. The collinearity between the determinants was assessed using pair-wise correlation, and those with absolute correlation coefficients above 0.7 were excluded from the models. During the multivariable logistic regression, variables with p≤ 0.05 were declared as significant determinants of modern contraceptive use in SSA.

Three models were fitted. Model 1 was a multivariable model adjustment for the HBM variables, and Model 2 adjusted for the SEM variables. In Model 3, all the statistically significant variables from Models 1 and 2 were fitted with the outcome variable to obtain the final determinants.

### Ethical approval

DHS surveys are approved by Inner City Fund(ICF) international and the ethical review boards of respective countries to ensure that the protocols comply with the United States Department of Health and Human Service regulations to protect human subjects[24]. This study involved secondary data analysis with no client contact.

## Results

### Demographic characteristics

The included surveys were conducted from 1990 to 2019, with 501,324 respondents across all the surveys. Most of the respondents were in the 15-24 age band (39.77%) and had up to primary education (32.44%). About two-thirds of them were married (63.58%) and dwelling in rural areas (62.5%), (table 2).

**Table 2.**
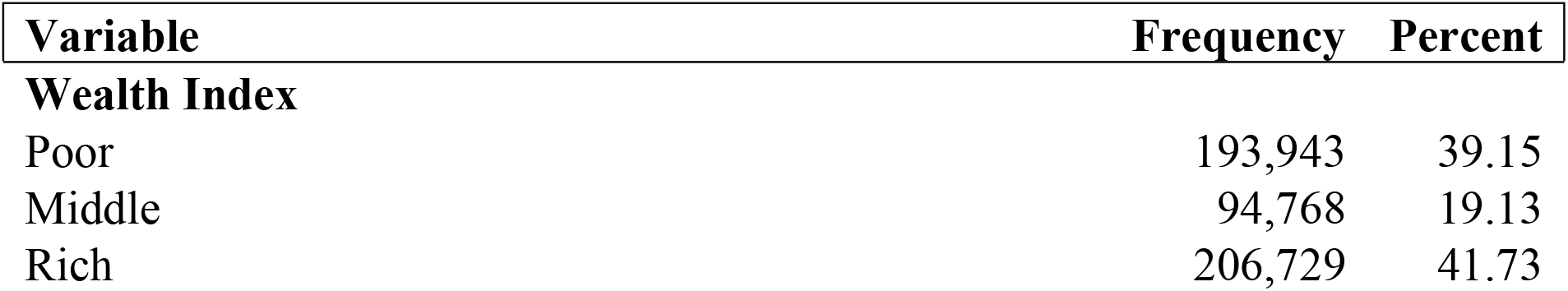
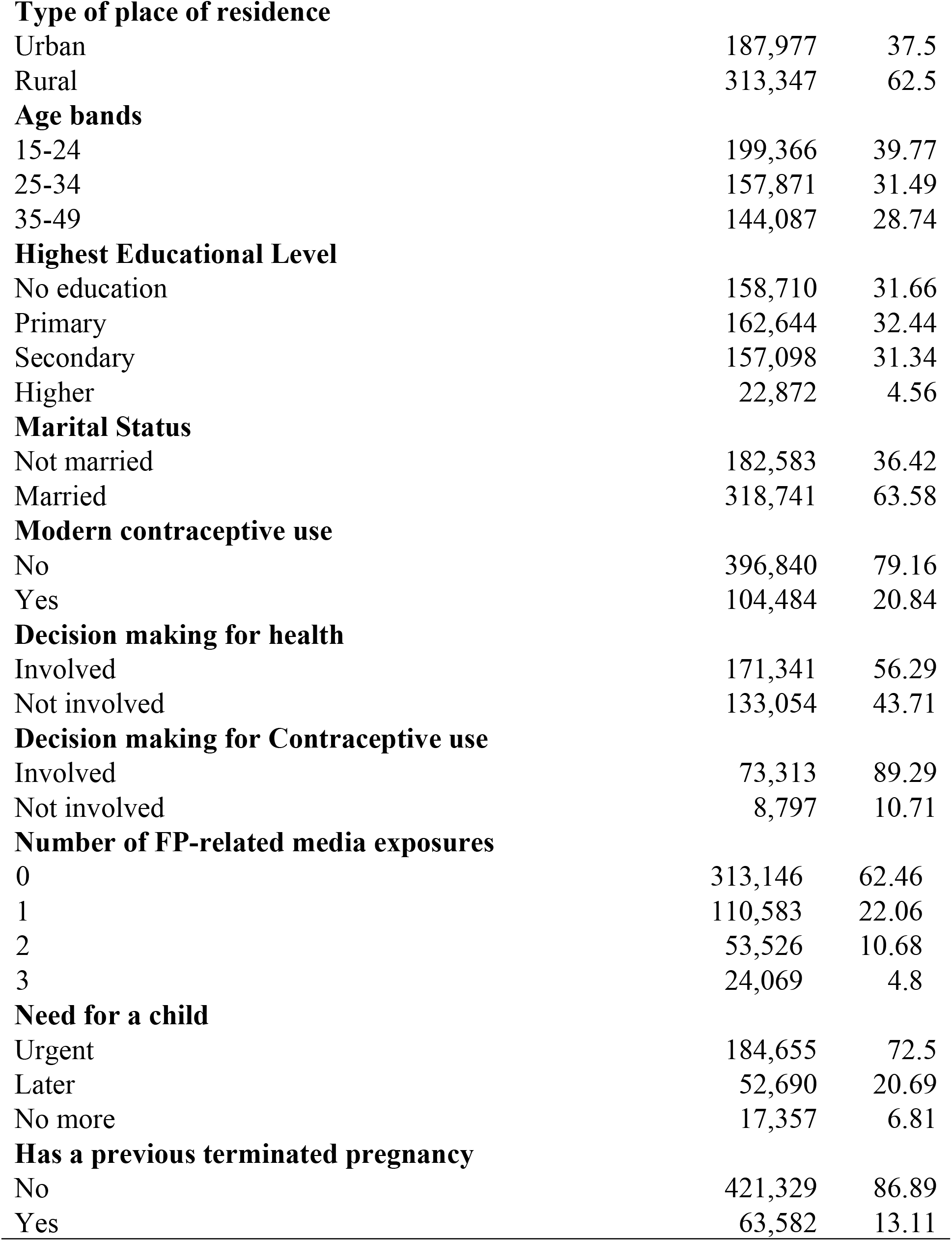
Socio-Demographic characteristics, modern contraceptive use, and distribution of modern contraceptive use determinants among respondents.

### Country-specific and pooled estimate of modern contraceptive use

The modern contraceptive coverage ranged between 3.47 % (95% CI: 2.92,4.02) in Central African Republic and 49.72% (95% CI:48.37,51.07) in Namibia. The pooled prevalence for the sub-region was 22.26 % (95% CI:17.91,26.60). Across the sub-regions, the Southern African countries had the highest pooled coverage (46.12%, 95% CI: 40.17, 51.26), and the lowest was within the West African sub-region (14.42%, 95% CI:11.42-17.41), (Fig 1).

### Determinants of modern contraceptive use

During the bivariate logistic regression, all the included variables in both the HBM and SEM were significantly associated with modern contraceptive use. Although being married increased the odds of modern contraceptive use (OR =1.68,95% CI:1.62, 1.75), it was not included in the modeling due to its colinearity with multiple variables. During the multivariable logistic regression in Models 1 and 2, ability to refuse sex (AOR=1.04,95% CI: 0.98,1.12), being in wealth index (Middle AOR=0.99,95% CI:0.87,1.14) and Rich (AOR=1.03, 95%CI: 0.89,1.21) and involvement in decision making for contraceptive use (AOR=1.05,95% CI: 0.92,1.21) were not significantly associated with modern contraceptive use so were excluded from Model 3. The variables that remained significant in both models are captured in table 3. All the variables in model 3, but urban residence (AOR=0.98,95% CI: 0.92,1.04), were significantly associated with modern contraceptive use.

**Table 3.**
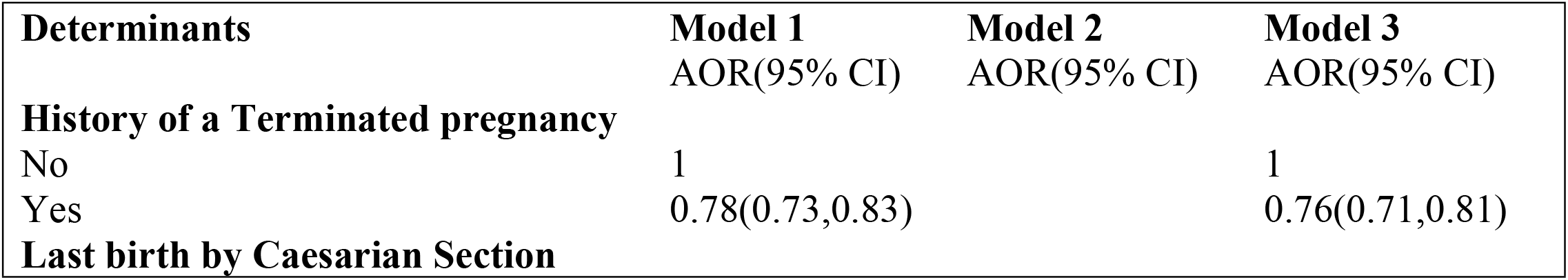
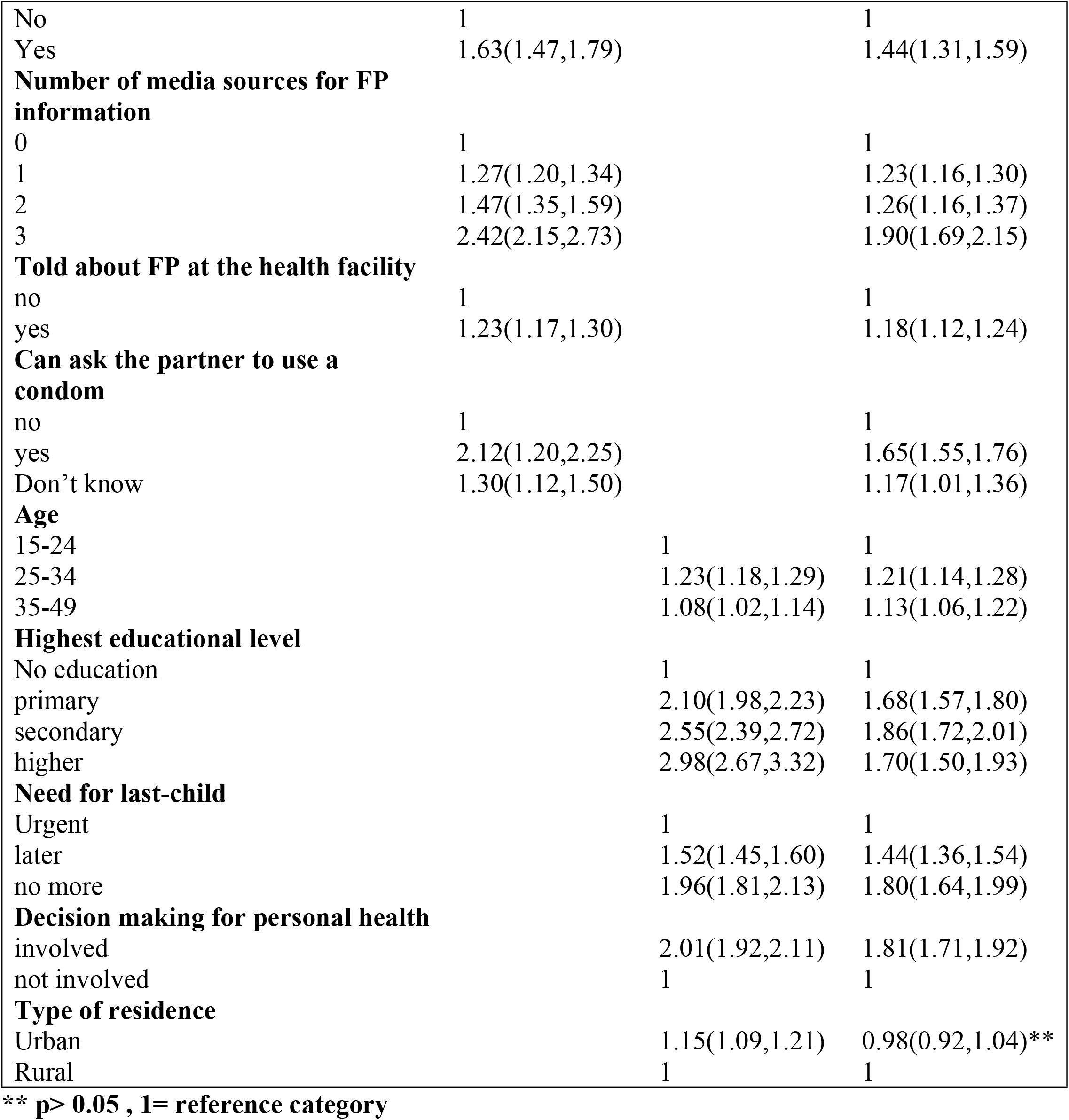
Multivariable multilevel logistic regression analysis of Health belief Model and Social-Ecological model variables associated with Modern contraceptive use in SSA.

Among the HBM variables, women whose last birth was by caesarian section were 1.44 times more likely to use a modern contraceptive than those who did not have a caesarian section (AOR=1.44, 95% CI:1.31,1.59). Compared to those who did not hear of family planning (FP) from any media outlet, a dose-dependent relationship was observed between modern contraceptive use and the number of media sources from which respondents received FP information. While hearing the information from 1 media source increased the odds by 23% (AOR=1.23,95% CI:1.16,1.30), hearing it from 2 sources increased it by 26%(AOR=1.26, 95% CI: 1.16,1.37) and it was increased by 90% for those who heard it from 3 sources (AOR=1.90, 95% CI: 1.69,2.15). Compared to those not told, being informed about family planning at the health facility increased the odds of modern contraceptive use by 18% (AOR=1.18, 95% CI:1.12,1.24).

The odds of modern contraceptive use increased by 65% among persons who could ask their partners to use a condom compared to those who could not (AOR=1.65, 95% CI: 1.55, 1.76). Even those who did not know whether they could ask their partner to use a condom were 1.17 times more likely to use a modern contraceptive than those who could not (AOR=1.17, 95% CI: 1.01, 1.36). However, having had a previous terminated pregnancy reduced the odds of modern contraceptive use by 24% (AOR=0.76, 95%CI: 0.71, 0.81).

On the SEM variables, there was a 21% increased likelihood among persons 25-34 years (AOR=1.21,95% CI:1.14,1.28) and a 13% increased odds among persons 35-49 (AOR=1.13,95% CI: 1.06,1.22) of modern contraceptive use compared to those 15-24 years. Compared to those who were not educated, having up to primary education increased the likelihood of modern contraceptive use by 68% (AOR=1.68, 95% CI=1.57.1.80). This likelihood increased to 86% among those who had up to secondary education (AOR=1.86, 95% CI: 1.72,2.01) and 70% among those schooled beyond the secondary level (AOR=1.70,95% CI: 1.5,1.93). Compared to those who needed the last child at the time of the survey, those who wanted the last child later were 1.44 times more likely to use modern contraceptives (AOR=1.44, 95% CI: 1.36,1.54), and those who needed no more children were 1.8 times more likely to use a modern contraceptive (AOR=1.8, 95% CI: 1.64,1.99). Being involved in decision-making concerning their own health increased the likelihood of modern contraceptive use by 81% compared to those not involved (AOR=1.81,95% CI:1.71,1.92).

## Discussion

The study sought to find the pooled prevalence of modern contraceptive coverage and its determinants among women in fertility age in SSA. The pooled prevalence was 22.26%. With respect to the determinants, women in fertility age (WIFA) who are more than 24 years and have a minimum of primary education were more likely to use modern contraceptives. Last birth by caesarian section, being exposed to family planning information on at least one media source or at the health facility, and the ability to negotiate condom use were also found to increase modern contraceptive use. WIFA who had no urgent need for a last child were more likely to use modern contraceptives than those who needed one urgently. Finally, those involved in decision-making concerning their health were more likely to use modern contraceptives than those not involved.

Being the second most populous and the region with the fastest-growing population, the low coverage of modern contraceptives is of significant concern because only Namibia nearly met the global SDG 3.7.1 thresholds of at least 50% modern contraceptive coverage among WIFA[26]. For the region to achieve its growth potential and harness all its resources for development, Aliyu calls for modern contraceptives to be vigorously promoted, not only for their demographic dividends but also for their socio-economic and health-related benefits that will accelerate the attainment of the sustainable development goals[27]. Evidence, however, needs to be used to guide the contraceptive promotion process, and the theory-based determinants identified in the study can serve as a good starting point.

The finding that exposure to family planning information in newspapers, on TV, or on radio increased the odds of modern contraceptive use in a dose-dependent fashion calls for a strong collaboration between health institutions, other key stakeholders, and media organizations in order to replicate the successful contribution media houses made towards family planning campaigns in Rwanda, Ethiopia, and Malawi across the SSA region[28]. With the majority of these women being on social media[29], there is the need to look beyond these traditional media outlets and add social marketing approaches, which are relatively cheaper and have demonstrated success in several campaigns within the region[30–32].

Although Boniface reported that some health workers hamper the use of contraceptives through their judgments of parity, religious beliefs, and other unfounded restrictions[33], evidence from this study adds to previous publications that demonstrated the role of health workers in increasing contraceptive coverage in the sub-region[34]. However, there have been calls for community health workers to be deployed to undertake these contraceptive promotion activities within communities as part of their routine work, rather than restricting them to health facilities[34].

The calls to empower WIFA towards achieving the SDGs[35–37] are further strengthened by the findings that those who could negotiate for condom use and those involved in decision-making concerning their health were more likely to use modern contraceptives. Aside from its direct effect on contraceptive use, Heera showed empowered women in urban slums to have lower rates of abortion. Though Prata and colleagues did not observe any relationship between empowerment and unintended pregnancies, they found empowered women to access antenatal care completely have lower maternal mortality rates and delayed sexual debut among their daughters[38,39].

While attempts are made to empower the WIFA in SSA, there is the need to prioritize those between 15 to 24 years who were found in this study to be less likely to use a modern contraceptive method. Contributing to this low likelihood are the cultural myths surrounding sex amongst young persons and country-specific regulations concerning the legal age of consent for sex and access to contraceptives[40–42]. These need to be considered in developing context-specific interventions for this particular age group. To empower them, the call by SDG 4 to “ensure inclusive and equitable quality education and promote lifelong learning opportunities for all” was found in this study to be very crucial in improving modern contraceptive use. There is a need for countries to ensure adherence to the provision of universal basic education, at least to the primary level, for all citizens[43].

The increased odds of modern contraceptive use among women whose last delivery was by caesarian in this study was observed by Bryant to be due to the education provided to them to delay conception for the uterine scar to heal well before the subsequent pregnancy and the introduction of long term methods(tubal ligation and IUCD) during the delivery process[44,45]. The findings that the urgent need for the last child decreased the odds of modern contraceptive use was observed by Coomson, who attributed it to the social desire to meet fertility needs and thus not only limited to the SSA region[46].

The decreased odds of modern contraceptive use among persons with a history of terminated pregnancy in the region might point to significant service delivery gaps because it increased contraceptive use in other contexts[47]. This gap is likely to be in providing comprehensive post-abortion care in the region. To resolve this, Corbett recommends ensuring the establishment and equipping of comprehensive one-stop-shop facilities to provide abortion services, making all forms of contraceptives available, and getting the whole package covered by health insurance[48]. Although urban residence has been shown to increase modern contraceptive use in several settings[49,50], its failure to significantly impact modern contraceptive use in the final model needs to be investigated further. However, it might be because most (62.5%) respondents were from rural dwellings.

### Recommendation

Countries in the region need to invest in developing context-specific interventions considering these determinants to help improve their contraceptive coverage and reduce the poor maternal and child health outcomes that result from unplanned pregnancies. Interventional studies need to be conducted in SSA to assess the effectiveness and cost-benefit ratio of interventions developed from these theory-based interventions in improving modern contraceptive use in the region.

### Strengths and limitations

The study’s strengths lie in using nationally representative data pooled together to obtain a large sample size, making the findings generalizable primarily to the included countries. The use of behavior change theories to objectively select determinants also increases the likelihood of recommendations impacting contraceptive use positively. However, a causal association cannot be established between modern contraceptive use and the determinants due to the cross-sectional nature of the survey. Not all the countries in the region had DHS data, so the findings may also not be representative of the entire SSA region.

## Conclusions

Modern contraceptive coverage in SSA is low. Age, educational status, past obstetric history, exposure to family planning information, ability to negotiate condom use or make personal health-related decisions, and the need for a child were the determinants for MC use in the region. To improve its coverage, interventions should be focused on empowering women through increased enrolment of girls in school, providing sexual and reproductive rights education, and increasing access to family planning information.

## Data Availability

The DHS dataset is freely available for use upon request at https://dhsprogram.com/data/available-datasets.cfm. After the request is approved, a de-identified dataset will be made available. The authors confirm they had no special access or privileges that others would not have.

## Acknowledgement

We thank Measure DHS for granting access to the data used for this study.

